# A Randomized, Controlled, Double Blind Clinical Study to Evaluate Use of Hydron Alkaline Ionised Water (HAIW) in Healthy Participants

**DOI:** 10.64898/2026.06.17.26355908

**Authors:** Vineet Kumar Malhotra, Sanjay Tamoli, Rahul Kalbhor, Sanjay Nipanikar, Ruby Dubey, Ritika Singhvi, Siddharth Sharma

## Abstract

**Background and Objectives:** Alkaline Ionized Water (AIW) is considered among the highest quality healthy drinking water worldwide and is widely discussed for its various health benefits. Hydron Alkaline Ionized Water (HAIW) is produced through electrolysis, resulting in a stable pH of approximately 9.5 with a negative Oxidation Reduction Potential (ORP), making it an antioxidant beverage. The objective of this study was to evaluate the safety of HAIW and its effects on digestion, sleep, energy, and overall quality of life in healthy participants compared to Packaged Drinking Water (PDW).

**Materials and Methods:** A randomized, controlled, double blind, prospective clinical study was conducted at Shivam Multi-specialty & Accident Care Centre Pvt. Ltd., Pune. A total of 24 healthy participants (21–40 years) were randomized in a 1:1 ratio to either HAIW Group (Group A, n=12) or Packaged Drinking Water Group (Group B, n=12), with equal gender distribution. Participants were hospitalized for 7 days and asked to consume at least 3 litres of the assigned water daily. Primary outcomes were safety-related laboratory parameters (CBC, LFT, RFT, Blood Sugar, ECG, Serum Electrolytes) and adverse event monitoring. Secondary outcomes included assessment of digestion (appetite, digestion, bowel habits), urine parameters, sleep quality, freshness after waking, fatigue (FSS), energy/stamina/strength, quality of life (WHO QOL BREF), and global assessment (CGI-I Scale).

**Results:** All 24 participants completed the study with no dropouts. Baseline demographics were comparable between the two groups. Assessment of primary safety-related laboratory parameters including CBC, liver function tests, renal function tests, blood sugar, ECG, and serum electrolytes showed non-significant change from baseline to 7 days and remained within normal limits in both groups, with non-significant difference between groups (p>0.05). HAIW showed significantly better improvement in appetite, digestion, and bowel habits from Day 2 onwards compared to PDW. Sleep quality and freshness after waking up showed significant improvement from Day 3 and Day 2 respectively in the HAIW and PDW group, with significantly better improvement in HAIW group. Fatigue scores showed significant reduction at Day 6 and 7 in both groups with non-significant difference between groups. A total of 5 adverse events were reported (3 in HAIW, 2 in PDW), all unrelated to study products and were mild in nature. Global assessment showed excellent to good overall safety and tolerability in both groups.

**Conclusion:** HAIW was well tolerated by all participants without any adverse effects. All laboratory safety parameters remained within normal range. HAIW demonstrated significant improvements in digestive function (appetite, digestion, bowel habits), sleep quality, and freshness after waking as compared to PDW. The study concludes that HAIW can be safely consumed. HAIW improves digestive and sleep-related functions.

## INTRODUCTION

Alkaline Ionized Water (AIW) is considered the highest level of healthy drinking water worldwide and is the most discussed functional beverage amongst researchers for its various health benefits. Alkaline Ionized Water is produced through electrolysis, as a result of which alkalinity is an inherent property of the water, unlike alkaline water produced by adding alkaline minerals. AIW thus has a stable pH value in the range of 8.5–9.5, is micro-clustered with a greater concentration of H+ ions (Hydrogen ions).

Alkaline water is deemed healthy for the body as it has a negative Oxidation Reduction Potential (ORP). ORP in water is a measure of whether the water oxidizes or reduces another substance inside the body. The more negative the ORP, the more anti-oxidizing is the water. The human body benefits from a negative ORP and alkaline environment for better physiological functions. This helps in restoring blood pH and provides faster hydration, thereby offering several health benefits, including anti-ageing, superior lubrication for muscles and joints, and enhanced immunity. Various Clinical trials have been conducted on AIW for its health benefits ^1–13^.

Some of the health benefits of Hydron AIW include:

- Faster hydration
- Anti-oxidant and anti-ageing capabilities
- Anti-hangover capabilities (post alcohol consumption)
- Increase in energy reserves
- Enhances immune system
- Improves cholesterol profile
- Ability to overcome acidity and constipation

**Table 1:** Difference between Different Water Types (HAIW, RO, Natural Alkaline Water, Artificially Made Alkaline Water)

| **Parameters** | **HAIW** | **RO Water** | **Natural Alkaline Water** | **Artificially Made Alkaline Water** |
| --- | --- | --- | --- | --- |
| **Manufacturing Process** | Electrolysis of water | Reverse osmosis; strips off almost all TDS including naturally found essential minerals | Alkalinity due to naturally occurring minerals; usually high in sodium, calcium and bicarbonates | Additives: Infusion of artificial minerals, bicarbonates to increase alkalinity |
| **pH Level** | Stable pH of ~9.5 | 6.5–8.5 (BIS standard) | 7–8.5 | Unstable pH of up to 9.5 |
| **Micro-clustered Water** | Yes (clusters of 4–6 molecules each) | No | No | No |
| **ORP (anti-oxidant potential)** | -300 to -1,000 mv (at time of dispensing) | No | No | No |
| **TDS (PPM, BIS standard)** | ~150 | ~200 (BIS Standard; due to remineralization) | 250–400 | 200 |
| **Product Classification** | Non-carbonated Water Based Beverage (pH above 8.5; BIS/FSSAI) | Packaged Drinking Water | Natural Alkaline Water | Non-carbonated Water Based Beverage (pH above 8.5; BIS/FSSAI) |

Various clinical trials have been conducted on AIW for its health benefits. To validate the claims of Hydron Alkaline Ionized Water, the present randomized, controlled, double blind clinical study was planned to evaluate the safety and health benefits of HAIW in healthy participants in comparison with Packaged Drinking Water (PDW).

## MATERIALS AND METHODS

### Study Design

A two arm, randomized, controlled, double blind, prospective clinical study was conducted. Participants were randomized in a 1:1 ratio to either the HAIW group or the PDW group. Identical opaque containers were used for both waters. The dispensing machine was operated by a third-party staff not involved in assessments. Both the investigator and participants were blinded to the group assignment. The total study duration was 7 days of inpatient study product consumption. There were four visit time points: Screening Visit (up to 3 days), Baseline Visit (Day 0), daily assessments during Days 0–7 (in hospital), and End of Study Visit (Day 7).

The study was conducted at Shivam Multi-specialty & Accident Care Centre Pvt. Ltd., Solapur-Pune Highway, Tarawadi, Phursungi, Pune, Maharashtra 412308. The study was approved by the Independent Ethics Committee, Mhaske Hospital and Research Centre, Hadapsar, Pune on 29th May 2024, and registered with CTRI (CTRI/2024/06/068585) on 07/06/2024.

### Study Participants

Healthy male and female participants between 21 to 40 years of age (both inclusive) were screened for eligibility. All participants provided written informed consent. For female participants of childbearing potential, a urine pregnancy test was performed at screening. Participants were required to have normal laboratory parameters (non-clinically significant) and to have no health-related signs and symptoms for which they required regular medication. Non-pregnant, non-lactating females were included. Participants willing to follow the study protocol and voluntarily sign informed consent were included.

Participants were excluded if they had known severe or chronic disease, active malignancy, significant cardiovascular event within 12 weeks prior to randomization, chronic contagious infectious diseases (HIV, Hepatitis B/C, active TB), active metabolic or gastrointestinal diseases, diabetes or hypertension, use of any investigational drug within 1 month, known hypersensitivity to alkaline water of PDW, pregnant or lactating, were currently participating in any other clinical study, or had any other condition deemed unsuitable by the investigator.

### Study Interventions

Test Product: Hydron Alkaline Ionized Water (HAIW) was prepared using electrolysis of water with a stable pH of approximately 9.5. HAIW was dispensed using Hydron AIW-ATM, a self-serving automated dispensing machine with a capacity of 500–5,000 LPD and output pH up to 9.5.

Control Product: Packaged Drinking Water (PDW) that is commercially available packaged drinking water brand.

Participants in both groups were hospitalized for 7 days and were provided standard diet and all necessary facilities (washrooms, bed, recreational and physical activities). During the hospital stay, participants were asked to consume at least 3 litres of the assigned water (HAIW or PDW) daily, in addition to water used in food and refreshments. Daily water consumption was recorded.

### Primary and Secondary Objectives

#### Primary Objectives

The primary objectives of the study were to assess safety-related laboratory parameters including Complete Blood Count (CBC), Liver Function Tests (LFT), Renal Function Tests (RFT), Blood Sugar, Serum Electrolytes, and ECG from baseline to the end of the study (7 days), and adverse events based on clinical signs and symptoms over the period of 7 days.

#### Secondary Objectives

The secondary objectives of the study were to comparatively assess changes in digestion-related symptoms including appetite, digestion, and bowel habits from baseline to end of the study and between the two groups; to comparatively assess change in urine output and related symptoms from baseline to end of the study and between the two groups; and to comparatively assess change in sleep quality from baseline to end of the study and between the two groups. Assessment of change in overall quality of life from baseline to end of the study. Additionally, the level of energy, stamina, and physical strength was assessed on a 7-point scale ranging from +3 (very satisfied) to −3 (very dissatisfied) from baseline to end of the study visit and between the two groups. Change in daytime fatigue was assessed using the Fatigue Severity Scale (FSS) from baseline to end of the study visit and between the two groups. Global evaluation for overall change was assessed on the CGI-I Scale as per the physician and participants at the end of the study and between the two groups, and global assessment of overall safety of the study products was performed as per the physician and participants at the end of the study and between the two groups.

### Statistical Methods

In-house statistician performed statistical analysis using Graphpad statistical software. Data describing quantitative measures were expressed as mean ± standard deviation. Qualitative variables were presented as counts and percentage. Comparison of variables representing categorical data was performed using t test and Chi-square test. All P values were reported based on two-sided significance, and all the statistical tests were interpreted at least up to 5% level of significance. A p-value of <0.05 was considered statistically significant. The values of the last visit were considered for final analysis for participants who did not complete the study schedule (Last Observation Carry Forward) for intent to treat analysis.

## RESULTS

### STUDY POPULATION

A total of 24 participants were screened for possible recruitment. There were no screen failures and all 24 participants were enrolled and randomized. 12 participants were randomized to HAIW Group (Group A) and 12 to PDW Group (Group B), with equal gender distribution (6 males and 6 females in each group). All 24 participants completed the study with no dropouts.

#### Study Consort

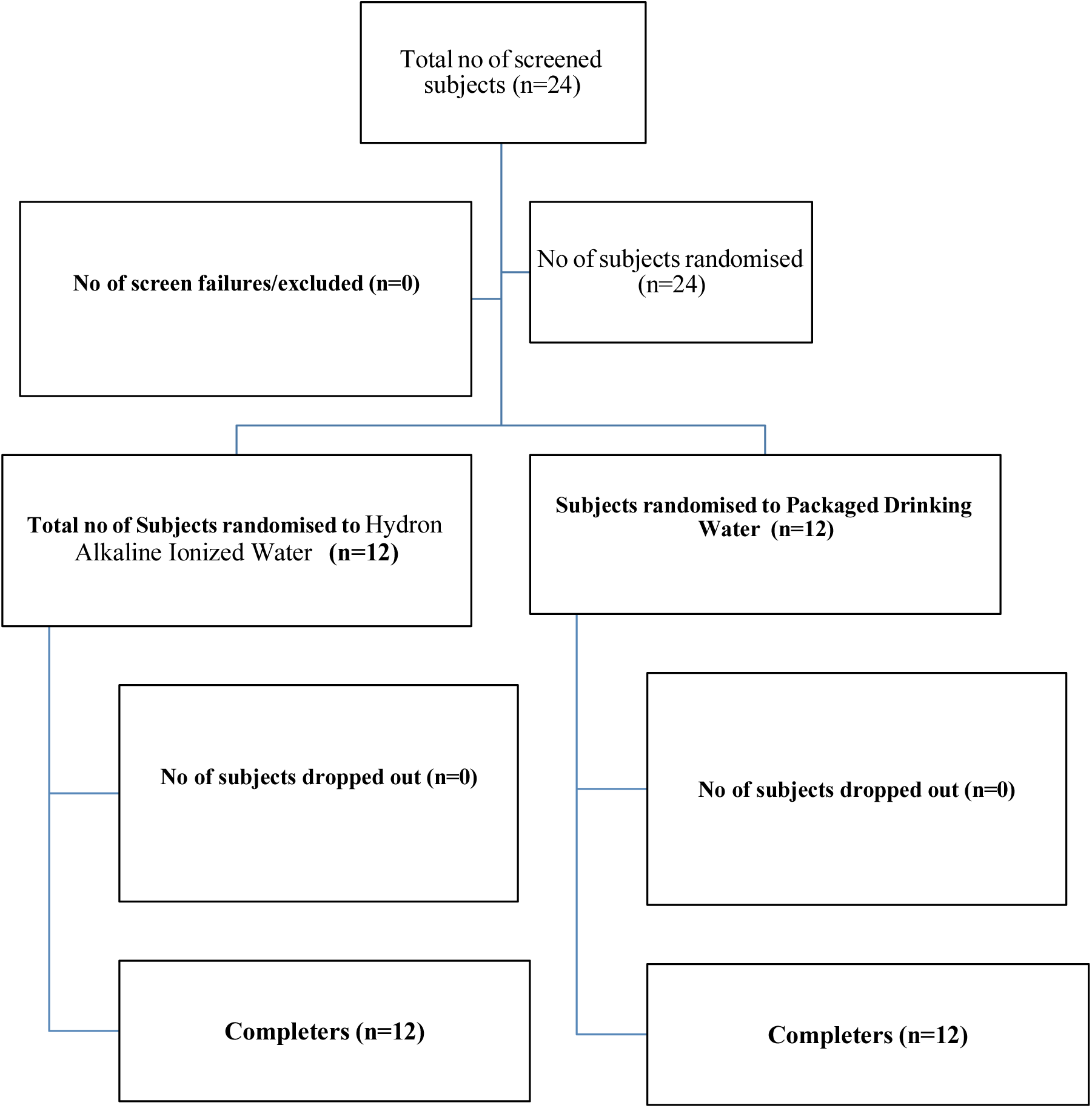

#### Baseline Demography

The baseline demography matched in the two groups with regards to gender, age, body weight and BMI. X Ray chest, ECG and USG findings were found to be normal and clinically non-significant in all the participants. Key characteristics are summarized in Table 2.

**Table 2:**
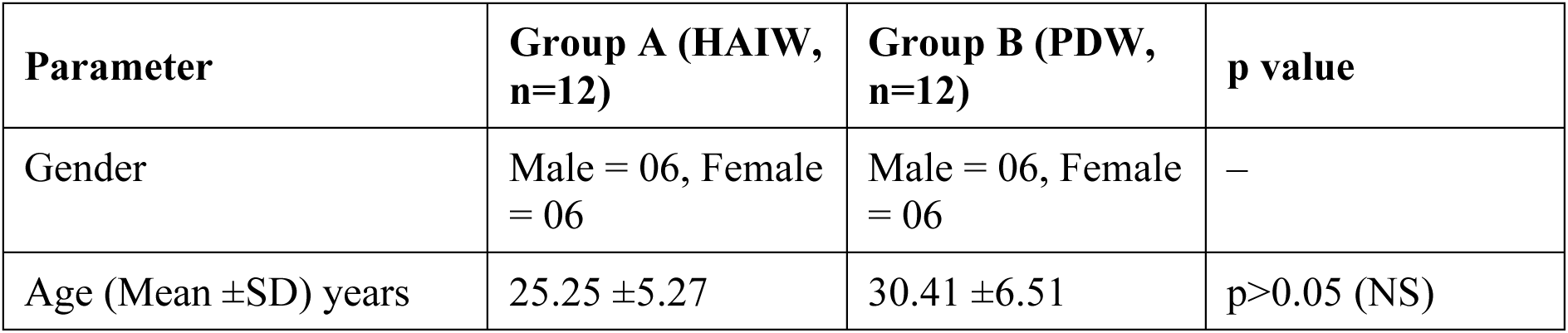
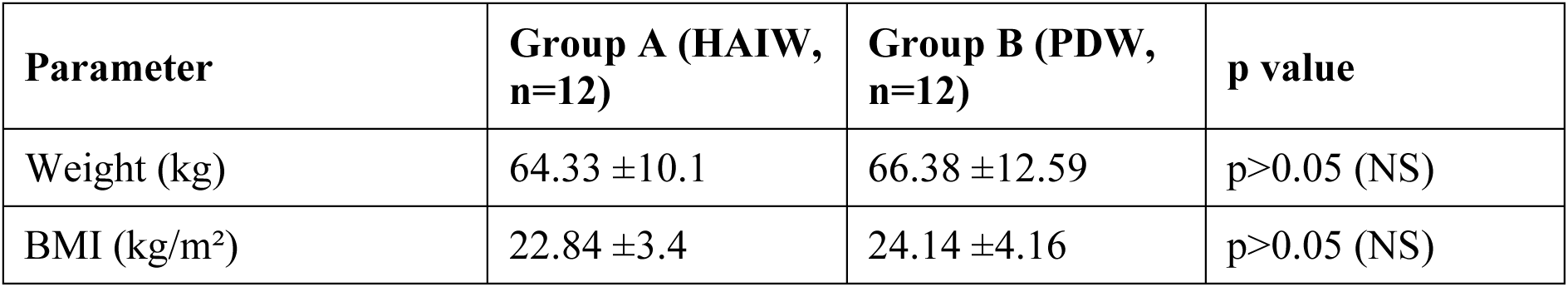
Assessment of Baseline Demography.

#### Assessment of Vital Parameters

Vital parameters including pulse, respiratory rate, temperature, SpO2, and blood pressure were observed to be within normal range at baseline and on all days during hospitalization through end of study in both groups, with non-significant difference between groups. Key summary values are shown in Table 3.

**Table 3:** Assessment of Vitals at Baseline and End of Study.

| Vitals | Group A – Baseline | Group A – End of Study | Group B – Baseline | Group B – End of Study |
| --- | --- | --- | --- | --- |
| Pulse (/min) | 82.16 ±10.41 | 80.83 ±7.10 | 79.66 ±9.19 | 82.25 ±5.73 |
| Respiratory Rate (/min) | 21.16 ±1.58 | 20.58 ±2.27 | 21.16 ±1.58 | 20.08 ±2.81 |
| Temperature (°F) | 97.9 ±0.39 | 97.76 ±0.44 | 97.75 ±0.43 | 97.96 ±0.40 |
| SpO<sub>2</sub> (%) | 97.83 ±0.71 | 97.91 ±0.51 | 97.75 ±0.86 | 97.66 ±0.88 |
| BP Systolic (mmHg) | 115.8 ±6.68 | 115.83 ±6.68 | 113.33 ±7.78 | 118.33 ±0.49 |
| BP Diastolic (mmHg) | 72.5 ±6.21 | 74.16 ±10.83 | 72.5 ±8.66 | 78.16 ±5.74 |
| p value within group | p>0.05 (NS) | – | p>0.05 (NS) | – |
| p value between groups | p>0.05 (NS) between the two groups |  |  |  |

##### Assessment of vitals – Pulse over a period of 7 days

Pulse rate over the study period of 7 days was observed to be within normal range and there was non-significant difference between the two groups at all the time points. Daily average pulse rate in the two groups has been mentioned in table 4.

**Table 4.** Assessment of Pulse (/min)

| Days | Group A | Group B |
| --- | --- | --- |
| Day 0 | 82.17 ±10.42 | 79.66 ±9.19 |
| Day 1 | 82.17 ±10.42 | 82.5 ±8.75 |
| Day 2 | 82.50 ±7.62 | 82.5 ±8.74 |
| **Day 3** | 78.83 ±6.15 | 78.5 ±10.61 |
| **Day 4** | 79.67 ±5.88 | 81.03 ±9.56 |
| **Day 5** | 78.58 ±6.93 | 77.91 ±4.77 |
| **Day 6** | 83.67 ±5.23 | 81.08 ±6.28 |
| **Day 7** | 80.83 ±7.11 | 84.25 ±5.73 |
| **p value within the group** | p>0.05(NS) from baseline to end of study | p>0.05(NS) from baseline to end of study |
| **p value between the groups** | p>(0.05) between the two groups |  |

##### Assessment of vitals – Respiration rate over a period of 7 days

Respiration rate over the study period of 7 days was observed to be within normal range and there was non-significant difference between the two groups at all the time points. Daily average pulse rate in the two groups has been mentioned in table 5.

**Table 5:** Assessment of Respiratory Rate (/min)

| Days | Group A | Group B |
| --- | --- | --- |
| Day 0 | 21.17 ±1.59 | 21.16 ±1.58 |
| Day 1 | 21.17 ±1.59 | 21.16 ±1.58 |
| Day 2 | 19.83 ±2.12 | 20.16 ±2.16 |
| Day 3 | 19.92 ±0.67 | 20.25 ±1.76 |
| Day 4 | 21.42 ±1.51 | 20.50 ±1.16 |
| Day 5 | 19.58 ±1.56 | 19.41 ±1.24 |
| Day 6 | 20.75 ±0.97 | 20.83 ±0.93 |
| Day 7 | 20.42 ±2.23 | 20.08 ±2.81 |
| p value within the group | p>0.05(NS) from baseline to end of study | p>0.05(NS) from baseline to end of study |
| p value between the groups | p>(0.05) between the two groups |  |

##### Assessment of vitals – Temperature over a period of 7 days

Body temperature over the study period of 7 days was observed to be within normal range and there was non-significant difference between the two groups at all the time points. Daily average pulse rate in the two groups has been mentioned in table 6.

**Table 6.**
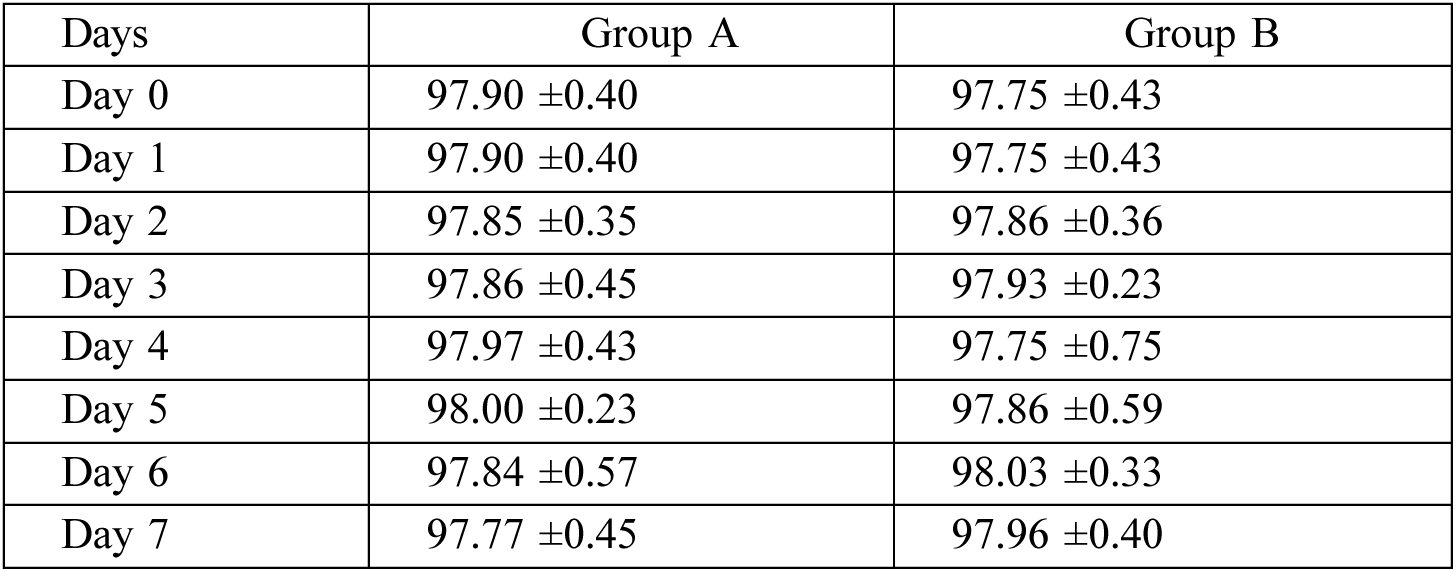
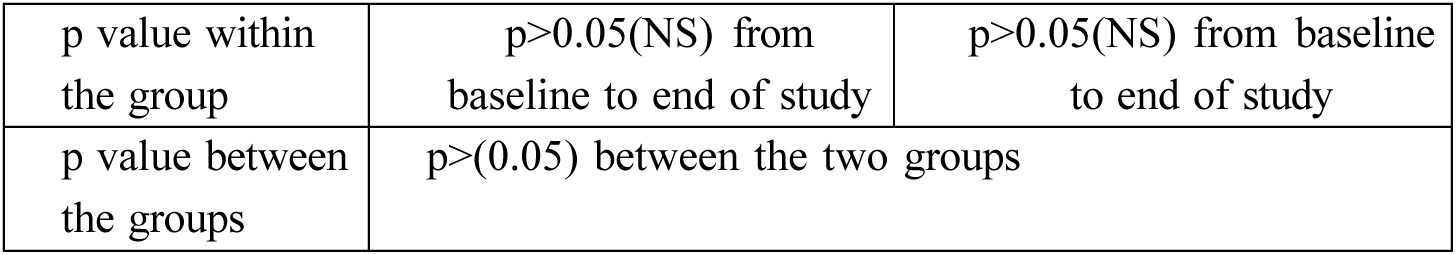
Assessment of Temperature (°F)

| Days | Group A | Group B |
| --- | --- | --- |
| Day 0 | 97.90 ±0.40 | 97.75 ±0.43 |
| Day 1 | 97.90 ±0.40 | 97.75 ±0.43 |
| Day 2 | 97.85 ±0.35 | 97.86 ±0.36 |
| Day 3 | 97.86 ±0.45 | 97.93 ±0.23 |
| Day 4 | 97.97 ±0.43 | 97.75 ±0.75 |
| Day 5 | 98.00 ±0.23 | 97.86 ±0.59 |
| Day 6 | 97.84 ±0.57 | 98.03 ±0.33 |
| Day 7 | 97.77 ±0.45 | 97.96 ±0.40 |
| p value within the group | p>0.05(NS) from baseline to end of study | p>0.05(NS) from baseline to end of study |
| p value between the groups | p>(0.05) between the two groups |  |

##### Assessment of vitals – SpO2 levels over a period of 7 days

SpO2 levels over the study period of 7 days was observed to be within normal range and there was non-significant difference between the two groups at all the time points. Daily average pulse rate in the two groups has been mentioned in table 7.

**Table 7.** Assessment of SpO2 (%)

| Days | Group A | Group B |
| --- | --- | --- |
| Day 0 | 97.85 ±0.71 | 97.75 ±0.86 |
| Day 1 | 97.85 ±0.71 | 97.75 ±0.86 |
| Day 2 | 97.77 ±0.97 | 97.98 ±0.74 |
| Day 3 | 97.78 ±0.73 | 98.16 ±0.57 |
| Day 4 | 98.75 ±0.45 | 98.16 ±0.93 |
| Day 5 | 98.00 ±0.85 | 97.33 ±0.98 |
| Day 6 | 98.17 ±0.58 | 97.75 ±0.86 |
| Day 7 | 97.42 ±1.78 | 97.66 ±0.88 |
| p value within the group | p>0.05(NS) from baseline to end of study | p>0.05(NS) from baseline to end of study |
| p value between the groups | p>(0.05) between the two groups |  |

##### Assessment of vitals – Blood Pressure (Systolic) over a period of 7 days

Blood Pressure (Systolic) over the study period of 7 days was observed to be within normal range and there was non-significant difference between the two groups at all the time points. Daily average pulse rate in the two groups has been mentioned in table 8.

**Table 8.** Assessment of BP Systolic (mm of Hg)

| Days | Group A | Group B |
| --- | --- | --- |
| Day 0 | 115.83 ±6.69 | 113.33 ±7.78 |
| Day 1 | 115.83 ±6.69 | 113.33 ±7.78 |
| Day 2 | 115.83 ±7.93 | 112.50 ±9.65 |
| Day 3 | 113.33 ±6.51 | 110.83 ±9.00 |
| Day 4 | 115.83 ±9.93 | 115.00 ±7.79 |
| Day 5 | 116.67 ±8.88 | 116.66 ±8.87 |
| Day 6 | 115.00 ±10.00 | 115.00 ±7.04 |
| Day 7 | 115.83 ±6.69 | 118.33 ±3.49 |
| p value within the group | p>0.05(NS) from baseline to end of study | p>0.05(NS) from baseline to end of study |
| p value between the groups | p>(0.05) between the two groups |  |

##### Assessment of vitals – Blood Pressure (Diastolic) over a period of 7 days

Blood Pressure (Diastolic) over the study period of 7 days was observed to be within normal range and there was non-significant difference between the two groups at all the time points. Daily average pulse rate in the two groups has been mentioned in table 9.

**Table 9.** Assessment of BP Diastolic (mm of Hg)

| Days | Group A | Group B |
| --- | --- | --- |
| Day 0 | 72.50 ±6.22 | 72.50 ±8.66 |
| Day 1 | 72.50 ±6.22 | 72.50 ±8.66 |
| Day 2 | 71.67 ±7.18 | 71.66 ±9.37 |
| Day 3 | 71.67 ±7.18 | 71.66 ±8.34 |
| Day 4 | 74.17 ±7.93 | 72.50 ±9.65 |
| Day 5 | 69.17 ±6.69 | 73.33 ±7.78 |
| Day 6 | 70.83 ±6.69 | 70.83 ±5.14 |
| Day 7 | 74.17 ±10.84 | 78.16 ±5.74 |
| p value within the group | p>0.05(NS) from baseline to end of study | p>0.05(NS) from baseline to end of study |
| p value between the groups | p>(0.05) between the two groups |  |

#### Assessment of Laboratory Parameters

Laboratory parameters including CBC, Blood Sugar (Fasting and Post Prandial), Liver Function Tests, Renal Function Tests, and Serum Electrolytes showed non-significant change from baseline to end of study in both groups. All values remained within normal range at baseline and end of study. There was non-significant difference between the two groups. Details shown in table 10 to 14.

**Table 10.** Assessment of CBC.

|  | Group A |  | Group B |  |
| --- | --- | --- | --- | --- |
| Tests | Baseline | End of Study | Baseline | End of Study |
| Hb (g/dL) | 13.76 ±1.81 | 13.63 ±1.72 | 13.95 ±2.65 | 13.90 ±2.52 |
| WBC(cells/μL) | 6155.83<br>±1614.73 | 5813.33<br>±946.09 | 6385.83<br>±1890.37 | 6737.50<br>±1696.42 |
| RBC (mill/c.mm) | 5.06 ±0.64 | 5.03 ±0.70 | 4.84 ±1.09 | 4.79 ±1.05 |
| Platelet Count (x10<sup>3</sup>/cumm) | 2.40 ±0.62 | 2.46 ±0.52 | 2.41 ±0.56 | 2.53 ±0.82 |
| ESR (mm/hr) | 7.41 ±7.69 | 7.58 ±7.91 | 8.50 ±6.74 | 10.58 ±9.88 |
| p value within the group | p>0.05(NS) from baseline to end of study |  | p>0.05(NS) from baseline to end of study |  |
| p value between the groups | p>(0.05) between the two groups at baseline and end of study |  |  |  |

**Table 11.**
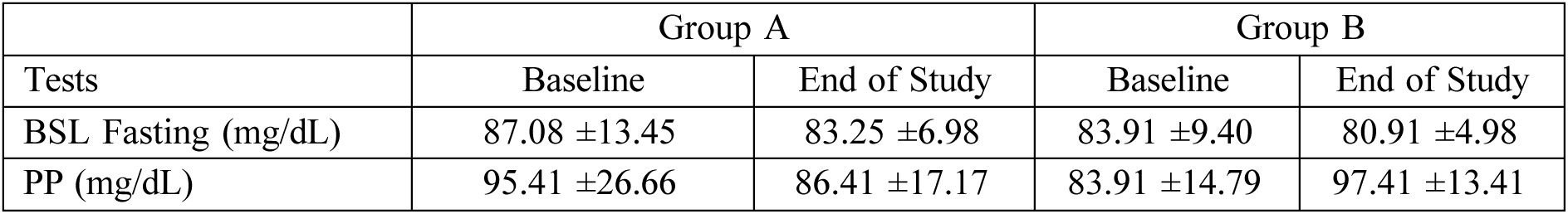
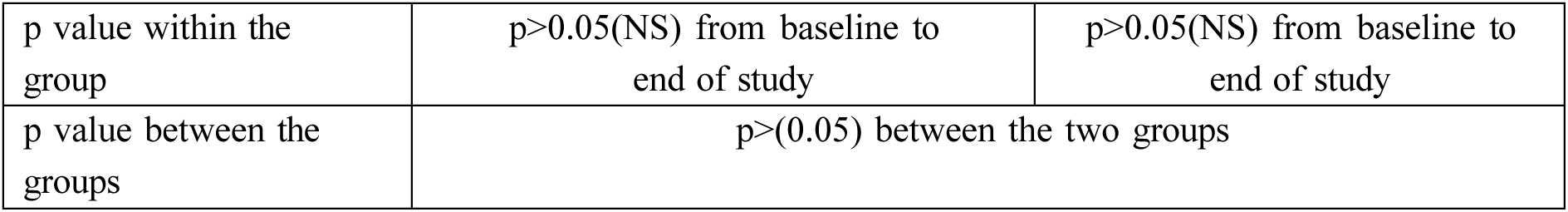
Assessment of BSF/PP.

| Tests | Group A |  | Group B |  |
| --- | --- | --- | --- | --- |
|  | Baseline | End of Study | Baseline | End of Study |
| BSL Fasting (mg/dL) | 87.08 ±13.45 | 83.25 ±6.98 | 83.91 ±9.40 | 80.91 ±4.98 |
| PP (mg/dL) | 95.41 ±26.66 | 86.41 ±17.17 | 83.91 ±14.79 | 97.41 ±13.41 |

|  |  |  |
| --- | --- | --- |
| p value within the group | p>0.05(NS) from baseline to end of study | p>0.05(NS) from baseline to end of study |
| p value between the groups | p>(0.05) between the two groups |  |

**Table 12:** Assessment of Liver Function Tests.

|  | Group A |  | Group B |  |
| --- | --- | --- | --- | --- |
| Tests | Baseline | End of Study | Baseline | End of Study |
| Sr. Bilirubin (mg/dL) | 0.64 ±0.19 | 0.63 ±0.16 | 0.67 ±0.26 | 0.53 ±0.17 |
| Dir. Bilirubin (mg/dL) | 0.20 ±0.6 | 0.20 ±0.03 | 0.20 ±0.05 | 0.15 ±0.03 |
| Indir. Bilirubin (mg/dL) | 0.44 ±0.13 | 0.43 ±0.13 | 0.47 ±0.21 | 0.38 ±0.14 |
| SGOT (U/L) | 22.15 ±3.98 | 23.16 ±5.92 | 22.37 ±2.72 | 21.00 ±3.99 |
| SGPT (U/L) | 18.44 ±5.45 | 20.00 ±6.72 | 18.75 ±4.46 | 17.57 ±8.76 |
| Sr. Protein (g/dL) | 7.52 ±0.39 | 7.37 ±0.35 | 7.69 ±0.38 | 7.39 ±0.22 |
| Sr. Albumin (g/dL) | 4.73 ±0.33 | 4.44 ±0.17 | 4.55 ±0.26 | 4.50 ±0.33 |
| Sr. Globulin (g/dL) | 2.79 ±0.42 | 2.9 ±0.29 | 3.13 ±0.24 | 2.88 ±0.35 |
| A/G Ratio | 1.74 ±0.42 | 1.57 ±0.18 | 1.47 ±0.13 | 1.59 ±0.29 |
| p value within the group | p>0.05(NS) from baseline to end of study |  | p>0.05(NS) from baseline to end of study |  |
| p value between the groups | p>(0.05) between the two groups |  |  |  |

**Table 13.**
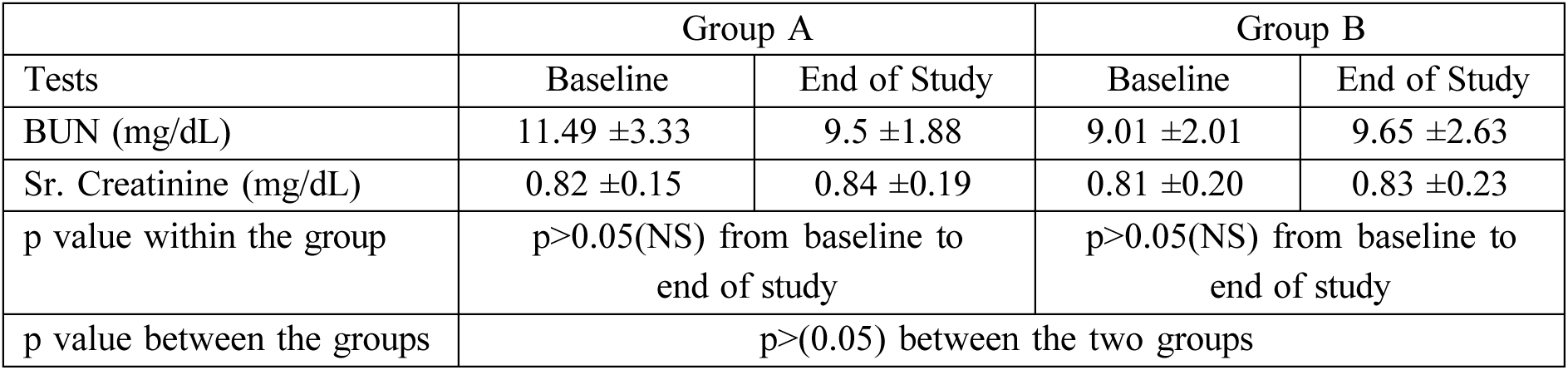
Assessment of RFT.

|  | Group A |  | Group B |  |
| --- | --- | --- | --- | --- |
| Tests | Baseline | End of Study | Baseline | End of Study |
| BUN (mg/dL) | 11.49 ±3.33 | 9.5 ±1.88 | 9.01 ±2.01 | 9.65 ±2.63 |
| Sr. Creatinine (mg/dL) | 0.82 ±0.15 | 0.84 ±0.19 | 0.81 ±0.20 | 0.83 ±0.23 |
| p value within the group | p>0.05(NS) from baseline to end of study |  | p>0.05(NS) from baseline to end of study |  |
| p value between the groups | p>(0.05) between the two groups |  |  |  |

**Table 14.** Assessment of Serum Electrolytes.

|  | Group A |  | Group B |  |
| --- | --- | --- | --- | --- |
| Tests | Baseline | End of Study | Baseline | End of Study |
| Sodium (mmol/L) | 142.16 ±2.20 | 142.25 ±1.76 | 141.5 ±2.90 | 141.91 ±1.37 |
| Potassium (mmol/L) | 4.18 ±0.36 | 4.22 ±0.43 | 4.30 ±2.50 | 4.29 ±0.52 |
| Chlorine (mmol/L) | 100.55 ±2.06 | 100.58 ±3.19 | 99.69 ±2.83 | 98.38 ±5.74 |
| p value within the group | p>0.05(NS) from baseline to end of study |  | p>0.05(NS) from baseline to end of study |  |
| p value between the groups | p>(0.05) between the two groups |  |  |  |

#### Secondary Efficacy Parameters

##### Assessment of symptoms related to digestive system

Participants were asked to rate their appetite, digestion, and bowel habits on a VAS scale of 0–100 (0 = very poor, 100 = excellent) on a daily basis.

##### Effect on Appetite

It was observed that there was a significant improvement in appetite score after the day 2 of consumption of HAIW which continued further till day 7. In PDW the same was observed after day 4 and continued till day 6 from baseline and on day 7 the same was found to be non-significant as compared to baseline. Between group analysis showed that there was a significantly better appetite improvement in HAIW group as compared to PDW from day 2 onwards which was observed till the end of the study. Details in table 15.

**Table 15.** Assessment of Appetite score over 7 days.

| Days | Group A | Group B | p value between the groups |
| --- | --- | --- | --- |
| Day 0 | 80.00 ±11.28 | 83.33 ±9.84 | p>0.05 (NS) |
| Day 1 | 80.00 ±11.28 | 83.33 ±9.84 | p>0.05 (NS) |
| Day 2 | 87.50 ±12.88* | 84.16 ±9.96 | p<0.05 (S) |
| Day 3 | 89.58 ±10.96* | 84.58 ±10.75 | p<0.05 (S) |
| Day 4 | 91.66 ±10.51* | 87.91 ±11.57* | p<0.05 (S) |
| Day 5 | 87.50 ±11.38* | 87.91 ±12.33* | p>0.05 (NS) |
| Day 6 | 89.16 ±13.11* | 87.50 ±10.55* | p<0.05 (S) |
| Day 7 | 91.25 ±13.50* | 83.33 ±8.61 | p<0.05 (S) |
|  | *p<0.05 from baseline (significant) | *p<0.05 from baseline (significant) |  |

##### Effect on Digestion

Digestion was assessed on VAS scale of 0-100 where 0 represented very poor digestion and 100 as excellent digestion. It was observed that there was a significant improvement in digestion score after the day 2 of consumption of HAIW which continued further till day 7. In PDW the same was observed on day 6 from baseline and on day 7 the same was found to be non-significant as compared to baseline. Between group analysis showed that there was a significantly better digestion improvement in HAIW group as compared to PDW from day 2 onwards which was observed till the end of the study (except on day 6). Details in table 16.

**Table 16.** Assessment of Digestion Score over a period of 7 days.

| Days | Group A | Group B | p value between the groups |
| --- | --- | --- | --- |
| Day 0 | 80.00 ±10.44 | 86.66 ±8.87 | p>0.05 (NS) |
| Day 1 | 80.00 ±10.44 | 86.66 ±8.87 | p>0.05 (NS) |
| Day 2 | 86.25 ±11.89* | 87.91 ±8.90 | p<0.05 (S) |
| Day 3 | 93.33 ±8.87* | 85.4 ±8.90 | p<0.05 (S) |
| Day 4 | 95.41 ±7.21* | 87.91 ±11.57 | p<0.05 (S) |
| Day 5 | 93.33 ±9.21* | 85.41 ±11.17 | p<0.05 (S) |
| Day 6 | 92.5 ±10.58* | 91.25 ±7.42* | p>0.05 (NS) |
| Day 7 | 91.66 ±10.29* | 88.33 ±9.37 | p<0.05 (S) |
|  | *p<0.05 from baseline (significant) | *p<0.05 from baseline (significant) |  |

##### Effect on Bowel Habits

Bowel habits were assessed on VAS scale of 0-100 where 0 represented poor bowel habits and 100 as excellent bowel habits. It was observed that there was a significant improvement in bowel habits score after the day 2 of consumption of HAIW which continued further till day 7. In PDW the same was observed on day 7 from baseline. Between group analysis showed that there was a significantly better bowel habit improvement in HAIW group as compared to PDW from day 2 onwards which was observed till the end of the study. Details in table 17.

**Table 17.** Assessment of Bowel Habits.

| Days | Group A | Group B | p value between the groups |
| --- | --- | --- | --- |
| Day 0 | 79.16 ±14.43 | 87.08 ±8.10 | p>0.05 (NS) |
| Day 1 | 79.16 ±14.43 | 87.08 ±8.10 | p>0.05 (NS) |
| Day 2 | 88.33 ±11.79* | 87.91 ±9.87 | p<0.05 (S) |
| Day 3 | 90.83 ±9.00* | 87.75 ±10.17 | p<0.05 (S) |
| Day 4 | 92.08 ±10.32* | 87.08 ±10.10 | p<0.05 (S) |
| Day 5 | 92.91 ±7.52* | 87.50 ±9.88 | p<0.05 (S) |
| Day 6 | 92.50 ±8.66* | 87.58 ±9.64 | p<0.05 (S) |
| Day 7 | 92.50 ±8.66* | 91.25 ±8.10* | p<0.05 (S) |
|  | *p<0.05 from baseline (significant) | *p<0.05 from baseline (significant) | - |

##### Effect on Bowel Frequency

Non-significant difference was observed from base line to day 7 on daily basis in both the study groups. Between the groups also showed non-significant difference between the two groups. Details in table 18.

**Table 18.** Assessment of Bowel Frequency (No. of Times)

| Days | Group A | Group B | p value between the groups |
| --- | --- | --- | --- |
| Day 0 | 1.08 ±0.28 | 1.33 ±0.65 | p>0.05 (NS) |
| Day 1 | 1.08 ±0.28 | 1.33 ±0.65 | p>0.05 (NS) |
| Day 2 | 1.41 ±0.66 | 1.33 ±0.77 | p>0.05 (NS) |
| Day 3 | 1.66 ±0.38 | 1.50 ±0.52 | p>0.05 (NS) |
| Day 4 | 1.25 ±0.45 | 1.33 ±0.65 | p>0.05 (NS) |
| Day 5 | 1.66 ±0.38 | 1.33 ±0.49 | p>0.05 (NS) |
| Day 6 | 1.33 ±0.49 | 1.33 ±0.65 | p>0.05 (NS) |
| Day 7 | 1.08 ±0.28 | 1.08 ±0.28 | p>0.05 (NS) |
|  | p>0.05 from baseline to all follow up visits within the group | p>0.05 from baseline to all follow up visits within the group |  |

##### Effect on consistency of stool

Non-significant difference was observed from base line to day 7 on daily basis in both the study groups on stool consistency as observed by Bristol scale. Between the groups also showed non-significant difference between the two groups. Details in table 19.

**Table 19.** Assessment of Bowel consistency on Bristol Stool Scale (For scoring refer scale)

| Days | Group A | Group B | p value between the groups |
| --- | --- | --- | --- |
| Day 0 | 3.58 ±0.51 | 3.75 ±0.86 | p>0.05 (NS) |
| Day 1 | 3.58 ±0.51 | 3.75 ±0.86 | p>0.05 (NS) |
| Day 2 | 3.66 ±0.49 | 3.66 ±0.65 | p>0.05 (NS) |
| Day 3 | 3.88 ±0.39 | 3.50 ±0.52 | p>0.05 (NS) |
| Day 4 | 3.75 ±0.45 | 3.75 ±0.45 | p>0.05 (NS) |
| Day 5 | 3.77 ±0.45 | 3.38 ±0.51 | p>0.05 (NS) |
| Day 6 | 3.75 ±0.45 | 3.75 ±0.62 | p>0.05 (NS) |
| Day 7 | 3.75 ±0.45 | 3.75 ±0.45 | p>0.05 (NS) |
|  | p>0.05 from baseline to all follow up visits within the group | p>0.05 from baseline to all follow up visits within the group |  |

#### Effect on Urine Parameters

Non-significant difference was observed from baseline to Day 7 on daily basis in both study groups on urine frequency. Between-group comparison also showed non-significant difference. Urine frequency data are shown in Table 20.

**Table 20.** Assessment of Urine Frequency.

| Days | Group A | Group B | p value between the groups |
| --- | --- | --- | --- |
| Day 0 | 7.41 ±2.19 | 7.91 ±3.20 | p>0.05 (NS) |
| Day 1 | 7.41 ±2.19 | 7.19 ±3.20 | p>0.05 (NS) |
| Day 2 | 7.16 ±2.08 | 8.00 ±2.89 | p>0.05 (NS) |
| Day 3 | 8.00 ±2.29 | 8.58 ±2.90 | p>0.05 (NS) |
| Day 4 | 7.91 ±2.27 | 8.66 ±3.22 | p>0.05 (NS) |
| Day 5 | 7.19 ±1.92 | 8.25 ±3.46 | p>0.05 (NS) |
| Day 6 | 8.33 ±2.26 | 8.50 ±3.37 | p>0.05 (NS) |
| Day 7 | 6.00 ±1.90 | 7.66 ±3.08 | p>0.05 (NS) |
|  | p>0.05 from baseline to all follow up visits within the group | p>0.05 from baseline to all follow up visits within the group |  |

#### Effect on Sleep

##### Total Sleep Time

Non-significant difference was observed from baseline to Day 7 on daily basis in both study groups on total sleep time. Between-group comparison also showed non-significant difference. Details in table 21.

**Table 21.** Assessment of Total Sleep time.

| Days | Group A | Group B | p value between the groups |
| --- | --- | --- | --- |
| Day 0 | 7.44 ±0.03 | 7.08 ±0.03 | p>0.05 (NS) |
| Day 1 | 7.44 ±0.03 | 7.08 ±0.03 | p>0.05 (NS) |
| Day 2 | 7.28 ±0.03 | 6.30 ±0.05 | p>0.05 (NS) |
| Day 3 | 7.39 ±0.05 | 7.17 ±0.04 | p>0.05 (NS) |
| Day 4 | 7.42 ±0.04 | 7.13 ±0.04 | p>0.05 (NS) |
| Day 5 | 7.40 ±0.06 | 6.50 ±0.04 | p>0.05 (NS) |
| Day 6 | 7.14 ±0.04 | 6.50 ±0.04 | p>0.05 (NS) |
|  | p>0.05 from baseline to all follow up visits within the group | p>0.05 from baseline to all follow up visits within the group |  |

##### Quality of Sleep

Assessment of sleep quality on VAS scale showed a significant improvement from baseline to Day 3 onwards, which continued till end of study in the HAIW group. Non-significant difference was observed in the PDW group on all days from baseline. Between-group assessment showed significantly better sleep quality from Day 3 onwards in HAIW group compared to PDW. Details in table 22.

**Table 22.** Assessment of Sleep quality.

| Days | Group A | Group B | p value between the groups |
| --- | --- | --- | --- |
| Day 0 | 91.66 ±8.34 | 90.83 ±9.00 | p>0.05 (NS) |
| Day 1 | 91.66 ±8.34 | 90.83 ±9.00 | p>0.05 (NS) |
| Day 2 | 93.75 ±7.72 | 89.58 ±9.64 | p>0.05 (NS) |
| Day 3 | 95.88 ±6.68* | 89.58 ±10.10 | p<0.05 (S) |
| Day 4 | 95.83 ±5.14* | 90.83 ±9.00 | p<0.05 (S) |
| Day 5 | 97.08 ±4.50* | 92.08 ±8.38 | p<0.05 (S) |
| Day 6 | 97.50±4.52* | 90.83 ±9.00 | p<0.05 (S) |
|  | *p<0.05 from baseline (significant) | p>0.05 from baseline for all days | - |

#### Freshness After Getting Up

Assessment of freshness after getting up showed a significant change from baseline in HAIW group after day 2 which continued further till end of the study. Analysis between the group also showed a significant difference in HAIW as compared to PDW after day 2 which was observed till the end of the study. Refer Table 23 for details.

**Table 23.** Assessment of freshness after getting up.

| Days | Group A | Group B | p value between the groups |
| --- | --- | --- | --- |
| Day 0 | 92.5 ±7.53 | 90.00 ±8.52 | p>0.05 (NS) |
| Day 1 | 92.5 ±7.53 | 90.00 ±8.52 | p>0.05 (NS) |
| Day 2 | 95.41 ±6.55* | 89.58 ±7.52 | p<0.05 (S) |
| Day 3 | 95.83 ±6.68* | 91.25 ±8.01 | p<0.05 (S) |
| Day 4 | 96.66 ±4.93* | 90.83 ±9.00 | p<0.05 (S) |
| Day 5 | 97.08 ±4.50* | 92.08 ±8.38 | p<0.05 (S) |
| Day 6 | 98.33 ±3.89* | 90.83 ±9.00 | p<0.05 (S) |
|  | *p<0.05 from baseline (significant) | p>0.05 from baseline for all days |  |

#### Assessment of Fatigue

Fatigue was assessed using the Fatigue Severity Scale (FSS) from baseline daily for 7 days. Significant reduction in fatigue was observed in both groups at Day 6 and Day 7. There was no significant difference between the groups at any time point. Details are shown in Table 24.

**Table 24:** Assessment of Fatigue Severity Scale.

| Days | Group A | Group B | p value between the groups |
| --- | --- | --- | --- |
| Day 0 | 16.25 ±4.67 | 17.16 ±5.37 | p>0.05 (NS) |
| Day 1 | 16.25 ±4.67 | 17.16 ±5.37 | p>0.05 (NS) |
| Day 2 | 16.00 ±4.32 | 16.91 ±4.90 | p>0.05 (NS) |
| Day 3 | 15.16 ±3.32 | 16.66 ±5.36 | p>0.05 (NS) |
| Day 4 | 15.16 ±3.27 | 16.83 ±5.42 | p>0.05 (NS) |
| Day 5 | 15.00 ±3.38 | 15.91 ±4.23 | p>0.05 (NS) |
| Day 6 | 14.83 ±2.97* | 15.33 ±3.82* | p>0.05 (NS) |
| Day 7 | 14.58 ±2.93* | 15.33±3.82* | p>0.05 (NS) |
|  | *p<0.05 from baseline (significant) | *p<0.05 from baseline (significant) |  |

#### Assessment of Energy, Strength and Stamina

Change in energy, stamina, and physical strength was assessed on a 7-point scale (+3 to −3) at baseline and Day 7. Non-significant difference was observed between the two groups. Results are shown in Table 25.

**Table 25:** Assessment of Energy, Strength and Stamina Levels.

| Score | Group A – Baseline | Group A – Day 7 | Group B – Baseline | Group B – Day 7 |
| --- | --- | --- | --- | --- |
| +3 (very satisfied) | 0 | 2 | 0 | 2 |
| +2 (somewhat satisfied) | 3 | 5 | 3 | 5 |
| +1 (a little satisfied) | 5 | 4 | 9 | 5 |
| 0 (neutral) | 4 | 1 | 0 | 0 |
| Dissatisfied | 0 | 0 | 0 | 0 |
| p value between the groups from baseline to day 7 p>0.05 – Non significant. |  |  |  |  |

#### Assessment of Quality of Life

Assessment of quality of life on WHO QOL BREF scale showed non-significant changes from baseline to end of study in both the groups. Between-group assessment also showed non-significant difference. It is noted that the WHO QOL scale is typically assessed after an interval of at least 4 weeks of intervention; the present study was only 7 days in duration, which may explain the non-significant changes. Details are shown in Table 26.

**Table 26:** Assessment of Quality of Life on WHO QOL Scale.

| Domain | Group A – Baseline | Group A – End of Study | Group B – Baseline | Group B – End of Study |
| --- | --- | --- | --- | --- |
| Q1 | 3.91 ±0.27 | 4.58 ±0.49 | 4.00 ±0.00 | 4.58 ±0.49 |
| Q2 | 4.00 ±0.40 | 3.91 ±0.49 | 4.00 ±0.57 | 4.16 ±0.37 |
| D1 (Physical) | 23.25 ±1.36 | 23.58 ±1.32 | 23.82 ±1.34 | 24.41 ±1.25 |
| D2 (Psychological) | 17.00 ±2.68 | 18.91 ±2.36 | 18.75 ±2.58 | 20.33 ±1.92 |
| D3 (Social) | 11.91 ±0.86 | 12.41 ±0.64 | 12.33 ±1.02 | 12.83 ±1.06 |
| D4 (Environmental) | 29.41 ±2.31 | 32.50 ±1.93 | 30.66 ±1.64 | 33.41 ±1.44 |

##### Assessment of adverse events

A total of 5 adverse events were reported in the study — 3 in the HAIW group (running nose: 2 days; stomach pain: 1 day; headache: 1 day) and 2 in the PDW group (headache: 1 day each in 2 separate participants). All events were of mild nature and none were related to the study product or study procedure. All episodes resolved within the study period and no event required stoppage of study product.

##### Global Assessment of Overall Change (CGI-I Scale)

It was observed that 7 participants reported of excellent overall change while 3 reported of good and 2 of fair overall change as per the investigator in HAIW group. In the PDW group of the 12 participants 3 reported of excellent overall change, 6 reported of good and 2 of fair overall change. One participant in the PDW group reported of no change. None of the participants in either of the two study groups reported of worsening of their condition with the use of study products.

##### Global Assessment of Overall Safety

All participants in both groups reported excellent to good overall safety. The same was confirmed by the investigators. Overall tolerability was rated as excellent to good by the majority of participants and investigators in both groups.

## DISCUSSION

The present study was conducted to evaluate the safety of Hydron Alkaline Ionized Water (HAIW) and its effects on digestion, sleep, energy, and overall quality of life in healthy participants compared to Packaged Drinking Water (PDW). All 24 participants completed the study with zero dropout rate, reflecting good tolerability and acceptability of the study product. Baseline demographics were well matched between the two groups, ensuring comparability of results. All safety-related laboratory parameters including CBC, liver function tests, renal function tests, blood sugar, ECG, and serum electrolytes showed non-significant change from baseline to 7 days and remained within normal limits in both the groups. No adverse events related to the study products were reported, confirming the excellent safety profile of HAIW. The most clinically notable findings were in digestive function and sleep quality. HAIW demonstrated significantly better improvement in appetite, digestion, and bowel habits from Day 2 onwards compared to PDW. This early onset of effect may be attributed to the alkaline nature and negative ORP of HAIW, which may facilitate better gastric environment, improved gut motility, and enhanced absorption. The micro-clustered structure of HAIW may also contribute to faster cellular hydration, positively impacting digestive processes. Sleep quality showed significant improvement from Day 3 onwards in the HAIW group with significant between-group differences, while freshness after waking showed significant improvement from Day 2 onwards. These findings suggest that enhanced hydration at the cellular level and antioxidant effects of HAIW may positively influence sleep architecture and recovery.

Fatigue as measured by the Fatigue Severity Scale showed significant reduction in both groups at Day 6 and 7, with non-significant difference between groups, suggesting that the improvement in fatigue may be partially attributable to the inpatient setting and structured lifestyle during hospitalization rather than exclusively to HAIW consumption. Quality of life on WHO QOL BREF scale showed non-significant changes, which is expected given the 7-day study duration — the WHO QOL instrument is typically validated for assessment after at least 4 weeks of intervention. A longer study duration may reveal significant quality of life improvements with HAIW.

## CONCLUSION

The study concludes that HAIW was well tolerated by all study participants without causing any adverse effects. All safety-related laboratory parameters showed no significant change over the period of 7 days, confirming the safety of HAIW for consumption. HAIW showed significant improvement in digestive function including appetite, digestion, and bowel habits as compared to Packaged Drinking Water, with effects observable from Day 2 of consumption. Significant improvement in sleep quality and freshness after waking up was also observed with the use of HAIW compared to PDW. All vital parameters remained within normal range in both groups throughout the study period. Urine parameters showed non-significant change from baseline to 7 days and between groups, indicating no adverse effect of HAIW on urinary function. Energy, strength, and stamina levels showed improvement in the both groups with non-significant between-group difference. Fatigue showed significant reduction in the both groups at Day 6 and 7. Based on the results of this study, HAIW can be safely consumed without causing any adverse effects and improves important physiological functions including digestion and sleep quality. These findings support the potential of Hydron Alkaline Ionized Water as a safe, health-promoting functional beverage for healthy adults.

## Data Availability

All data produced in the present study are available upon reasonable request to the authors

## ACKNOWLEDGEMENTS

The authors acknowledge the contributions of all study participants, without whom this research would not have been possible. We extend our sincere gratitude to the sponsor, Hydron Alkaline Aqua Pvt. Ltd., Mumbai, India, for funding and supporting this clinical study. We also thank the Institutional Ethics Committees of study sites for their review and approval of the study protocol. We extend our sincere gratitude to the investigators and clinical staff at Shivam Multispeciality Hospital, Maharashtra for their dedication and support throughout the conduct of this study. Special thanks to the team at Target Institute of Medical Education & Research, for their contributions to study coordination and data management.

